# The prevalence, incidence, prognosis and risk factors for depression and anxiety in a UK cohort during the COVID-19 pandemic

**DOI:** 10.1101/2021.06.11.21258750

**Authors:** Ru Jia, Kieran Ayling, Trudie Chalder, Adam Massey, Norina Gasteiger, Elizabeth Broadbent, Carol Coupland, Kavita Vedhara

## Abstract

**Background:** The COVID-19 pandemic had profound immediate impacts on population mental health. However, in whom the effects may be prolonged is less clear.

**Aims:** To investigate the prevalence, incidence, prognosis, and risk factors for depression and anxiety reported in a UK cohort over three distinct periods in the pandemic in 2020.

**Method:** An online survey was distributed to a UK community cohort (n=3097) at three points: April (baseline), July-September (T2) and November-December (T3). Participants completed validated measures of depression and anxiety on each occasion and we prospectively explored the role of socio-demographic factors and psychological factors (loneliness, positive mood, perceived risk of and worry about COVID-19) as risk factors.

**Results:** Depression (PHQ-9 means - baseline: 7.69, T2: 5.53, T3: 6.06) and anxiety scores (GAD-7 means -baseline: 6.59, T2: 4.60, T3: 4.98) were considerably greater than pre-pandemic population norms. Women reported greater depression and anxiety than men. Being younger, having prior mental health disorders, more negative life events due to COVID-19, as well as greater loneliness and lower positive mood at baseline were significant predictors of poorer mental health outcomes.

**Conclusion:** The negative impact of the COVID-19 pandemic on mental health has persisted to some degree. Younger people and individuals with prior mental health disorders were at greatest risk. Easing of restrictions might bring the opportunity for a return to social interaction, which could mitigate the risk factors of loneliness and positive mood.

## Introduction

### Background and aims

The COVID-19 (SARS-Cov-2, 2019) pandemic has resulted in unprecedented disruptions to people’s daily lives, health care provision and the economy. There is a growing body of literature reporting evidence of a rapid and significant deterioration in the United Kingdom (UK) population’s mental health which occurred within weeks of the first national lockdown,(1-3) a pattern repeated in many countries.(4) However, the nature of the pandemic has changed over time, with levels of infection and mortality fluctuating, which in turn have precipitated changes in social restrictions. We describe here the prevalence, incidence, and prognosis of mental health difficulties reported in a UK cohort established early in the pandemic over three distinct periods in 2020. We aimed to explore: (1) how social restrictions in the UK at three key stages (first lockdown/Time 1 (baseline), eased restrictions/Time 2, increased restrictions/Time 3) impacted on anxiety and depression; (2) the socio-demographic and psychological factors at baseline that predicted anxiety and depression at Time 3 and (3) socio-demographic and psychological factors associated with the incidence and prognosis of depression and anxiety cases over time.

### Known and unknown from the evidence

The COVID-19 pandemic can be characterised as a chronic stressor,(5) in that it has now impacted most peoples’ lives for more than a year. There is no clear end in sight, and it is both unpredictable and largely uncontrollable at the level of the individual. In the UK, the trajectory of the pandemic in 2020 had several key phases. It commenced with the first national lockdown (23^rd^ March 2020) when people were instructed to stay at home and schools were closed. Aside from a small number of exceptions, for most of the UK, the lockdown was gradually eased from 11^th^ May 2020(6) with people allowed to meet others from outside their household with reopening of schools, hospitality and retail venues. This continued to early September. However, from September 2020 the number of areas in which local restrictions were tightened began to increase and the spiralling number of infections and deaths led inexorably to a second lockdown in November 2020 with many and fluctuating restrictions throughout December. One of the considerations of public health policy regarding the changes in social restrictions, was, and continues to be, its impact on mental health.(7) It is, therefore, important to examine whether mental health did indeed improve in response to eased restrictions and subsequently if a resumption of restrictions precipitated a deterioration. Evidence from longitudinal studies with large UK cohorts suggests that levels of anxiety and depression, for example, improved during summer 2020,(2, 8) but less is known about the impact of the autumn/winter lockdown. Beyond a simple description of how mental health has fluctuated in response to social restrictions, it is also of interest to examine whether the characteristics associated with mental health difficulties at the start of the pandemic remained consistent over time. For example, several studies demonstrated that young people and women were at greater risk of psychological distress early in the pandemic.(2, 8, 9) In addition to these demographic predictors, we and others have reported that greater perceived risk of COVID-19, worry about contracting COVID-19, loneliness and reduced positive mood were also associated with greater depression and anxiety during lockdowns in different countries and regions.(1, 9, 10) Finally, it is also relevant to examine the factors that predict how individuals’ mental health changed in response to the pandemic. The seemingly sudden and rapid deterioration in mental health for large swathes of the population was perhaps not unexpected considering that the pandemic was initially a novel and unprecedented experience for most people. However, stress and coping theory(5) would lead us to expect that some people will have been able to adjust to the challenges of the pandemic through identifying and implementing effective coping strategies.(11) Previous work from the Severe Acute Respiratory Syndrome (SARS) epidemic suggested that less SARS-related worry and greater social support were protective factors against subsequent mental health difficulties.(12) We report on these issues here by presenting analyses from a longitudinal community cohort from the UK.

## Methods

### Recruitment and eligibility

The authors assert that all procedures contributing to this work comply with the ethical standards of the relevant national and institutional committees on human experimentation and with the Helsinki Declaration of 1975, as revised in 2008. The University of Nottingham (Deleted for blind review) Faculty of Medicine and Health Sciences (ref: 506-2003) and the NHS Health Research Authority (ref: 20/HRA/1858) approved all study procedures. Recruitment processes were reported previously.(1) In short, participants were recruited in the community through a social and mainstream media campaign between the third and 30^th^ April 2020. NHS organisations were also approached to promote the research through their routine communications. Potential participants were directed to the study website (www.covidstressstudy.co.uk) through which they accessed the information sheet, consent form and online survey.

Eligible participants were aged 18 years and over; able to give informed consent; able to read English; residing in the UK at the time of completing the survey and able to provide a sample of hair at least one centimetre long. The latter was collected for the determination of the stress biomarker cortisol, which will be the subject of future manuscripts.

### Procedures

Consenting participants completed an online survey implemented through JISC Online Survey (https://www.onlinesurveys.ac.uk/). We administrated three periods of data collection: baseline between 3/4/20 and 30/4/20 (national lockdown), Time 2 between 1/7/20 and 21/9/20 (eased restrictions) and Time 3 between 11/11/20 and 31/12/20 (increased restrictions including four weeks of lockdown).

Participants who completed the baseline survey were invited by email to complete the survey again at Time 2 and Time 3. Socio-demographic factors (age, gender, ethnicity, keyworker status, being in a recognised COVID-19 risk category, living alone or with others) were collected at baseline. The following psychological measures were collected at all time points: anxiety (7-item Generalized Anxiety Disorder Scale, GAD-7, α=0.88)(13) and depression (Patient Health Questionnaire, PHQ-9, α=0.92).(14) The psycho-social factors we assessed were positive mood (Scale of Positive and Negative Experience-Positive, SPANE-P, α=0.94),(15) worry about contracting COVID-19, perceived loneliness and risk of COVID-19, details of which are reported elsewhere.(1) In addition, at Time 2 we asked participants whether they had prior mental health disorders and at Time 3, we asked whether participants had experienced any negative/positive life events due to COVID-19 (based on a brief checklist of events). Each event was scored for one and negative and positive events were totalled and scored separately (for item details see Supplementary Appendix S1)

### Statistical analysis

We first summarised the outcome variables (depression and anxiety scores) and participant characteristics with appropriate descriptive statistics and examined histograms and scatterplots for normality. Comparisons with pre-pandemic normative values were made using independent samples t-tests. Examination of histograms indicated that both depression and anxiety scores deviated from a normal distribution, however transformations or non-parametric tests were not suitable for these comparisons as only summary statistics (not individual-level data) were available for normative data. While t-tests are robust to deviations from normality especially when sample sizes are large,(16) results of these specific tests should be interpreted with caution. Depression and anxiety were also categorised based on original cut-offs.(13, 14)

Comparisons of mental health outcomes at three time points were made using repeated measures ANOVA. We conducted multivariable linear regression models to explore the independent relationships of socio-demographic factors (age, gender, ethnicity, keyworker status, prior mental health disorders, living alone, being in a recognised COVID-19 risk group, experience of positive/negative life events) and baseline psychological factors (perceived loneliness, perceived risk of COVID-19, positive mood, COVID-19 worry), with depression and anxiety scores at Time 3. The variable assessing COVID-19 worry was treated as a categorical variable in all models, with “occasional worry” treated as the reference value as this was the most common response. Assumptions of linear regression (normality and homoscedasticity of residuals, linearity with continuous variables) and presence of outliers were assessed graphically. Square root transformations were used for depression and anxiety scores to satisfy assumptions.

We conducted sensitivity analysis by using Multiple Imputation (MI) with chained equations to impute values for each variable with missing values (age, gender, ethnicity, prior mental health disorders, positive/negative life events, depression and anxiety at both Time 2 and 3). This approach is suitable for longitudinal data.(17) We generated 70 imputed datasets. Multivariable regression models predicting depression and anxiety scores at Time 3 were built with MI datasets, and estimates were combined using Rubin’s rules. Perceived risk was not significant in the main analyses hence was excluded from this sensitivity analysis.

To examine predictors of incidence and prognosis of depression and anxiety, we dichotomised depression and anxiety outcomes according to established cut-offs for ‘caseness’(18) where levels of symptoms reached the thresholds for high intensity psychological support (PHQ-9 score ≥ 10, GAD-7 score ≥ 8) in the NHS. Cochran’s Q tests were conducted to examine the differences in the proportions of depression and anxiety cases over time. Individuals who were not classified as cases of depression or anxiety at baseline but became cases at Time 2 or 3 were classified as incident depression or anxiety cases. Individuals who were classified as cases for depression or anxiety at baseline but subsequently became non-cases at Time 2 or 3 were further classified as remission of depression or anxiety cases. We used logistic regression to estimate odds ratios (ORs) with 95% confidence intervals for associations with incidence and remission of depression and anxiety cases at Time 3 using demographic and psychological factors at baseline, relative to no change of case status. Demographic and psychological factors that were significantly associated with depression or anxiety in the previous multivariable linear regression models were all included in the logistic regression analysis.

Statistical analyses were performed using STATA (version 16).

### Role of sponsor

The study sponsor did not play a role in the study design, collection; analysis, and interpretation of data; in the writing of the report; or in the decision to submit the paper for publication.

## Results

### Cohort characteristics

At baseline, 3097 participants completed the survey. Forty-five percent (n=1385) of this cohort returned the follow-up survey at Time 2 and 35% (n=1087) at Time 3. Twenty-eight percent (n=881) of the baseline respondents completed all three surveys. Three participants left the UK before Time 3 hence were removed from analyses. This resulted in a final cohort of 878 UK-dwelling participants who completed all three surveys (completers). Demographic and baseline mental health characteristics of the completers and non-completers of all three surveys (drop-outs) are presented in Table 1.

**Table 1:** Baseline characteristics in completers and drop-outs.

|  | Completers | Drop-outs |
| --- | --- | --- |
|  | n (%) | n (%) |
| **N** | 878 (28.4%) | 2216 (71.6%) |
| **Gender** |  |  |
| Male | 123 (14.0%) | 353 (15.9%) |
| Female | 754 (85.4%) | 1861 (84.0%) |
| Prefer not to say | 1 (0.1%) | 2 (0.1%) |
| **Age (mean, SD)\*** | 49.7 (15.0) | 42.6 (14.5) |
| **Age groups (years)** |  |  |
| 18-24 | 49 (5.6%) | 313 (14.1%) |
| 25-34 | 117 (13.3%) | 410 (18.5%) |
| 35-44 | 147 (16.7%) | 490 (22.1%) |
| 45-54 | 193 (22.0%) | 497 (22.5%) |
| 55-64 | 218 (24.8%) | 352 (15.9%) |
| 65-74 | 129 (14.7%) | 128 (5.8%) |
| ≥75 | 25 (2.9%) | 24 (1.1%) |
| **Ethnicity\*** |  |  |
| White – British, Irish, other | 826 (94.1%) | 1967 (88.9%) |
| BAME background | 51 (5.8%) | 245 (11.1%) |
| **Keyworker status \*** |  |  |
| Keyworker | 354 (40.3%) | 1204 (54.3%) |
| Not a keyworker | 524 (59.7%) | 1012 (45.7%) |
| **COVID-19 risk groups\*** |  |  |
| Most at risk (e.g. suffering from advanced cancer, severe asthma/COPD, etc.) | 25 (2.9%) | 96 (4.3%) |
| At increased risk (e.g., being pregnant, aged over 70) | 180 (20.5%) | 348 (15.7%) |
| Not at-risk | 673 (76.7%) | 1772 (80.0%) |
| **Living alone (or with others) \*** |  |  |
| Living alone | 134 (15.3%) | 271 (12.2%) |
| Living with others | 744 (84.7%) | 1945 (87.8%) |
| **Depression (mean, SD)\*** | 5.96 (5.2) | 8.37 (6.2) |
| **Anxiety (mean, SD)\*** | 5.15 (5.0) | 7.16 (5.7) |
| **Loneliness (mean, SD)\*** | 3.33 (2.5) | 4.1 (2.8) |
| **Positive mood (mean, SD)\*** | 20.08 (4.9) | 18.6 (5.1) |
| **Perceived risk of COVID-19 (mean, SD)\*** | 3.93 (1.87) | 4.9 (2.3) |
| **COVID-19 Worry\*** |  |  |
| No worry (n, %) | 191 (21.8%) | 359 (16.2%) |
| Occasional worry (n, %) | 626 (71.3%) | 1443 (65.1%) |
| Much worry (n, %) | 50 (5.7%) | 311 (14.0%) |
| Most worry (n, %) | 11 (1.3%) | 103 (4.7%) |

\* Significant difference between completers and drop-outs.

Significant differences in demographic and baseline mental health characteristics were found between completers and drop-outs. Specifically, individuals who were younger (*p*<.001), from ethnic minority backgrounds (*p*<.001), keyworkers (*p*<.001), not in a COVID-19 risk group (*p*=.002), and living with others (*p=*.02) were more likely to drop out from the study. Participants with poorer mental health characteristics at baseline were also more likely to drop out from the study. This included those with higher levels of depression (square-root transformed mean: 2.63 vs. 2.14, *p*<.001), higher levels of anxiety (square-root transformed mean: 2.38 vs. 1.92, *p*<.001), greater loneliness (mean: 4.06 vs. 3.33, *p*<.001), lower positive mood (mean: 18.56 vs. 20.08, *p*<.001), and more worry about getting COVID-19 (*p*=0.002).

### Depression and anxiety over time

Mean levels of depression and anxiety in the whole cohort at each time point are presented in Figure 1. The overall mean values for depression and anxiety were significantly higher than previously reported population norms,(19, 20) at all three time points (all *p*<.001). Female participants reported significantly higher levels of both depression and anxiety than male participants across time (depression (baseline: *p*<.001, Time 2: *p*<.001, Time 3: *p*=.001), anxiety (baseline: *p*<.001, Time 2: *p*<.001, Time 3: *p*=.002). The mean depression and anxiety scores for both genders were also significantly higher than their respective population norms (all *p*<.001).

**Figure 1.**
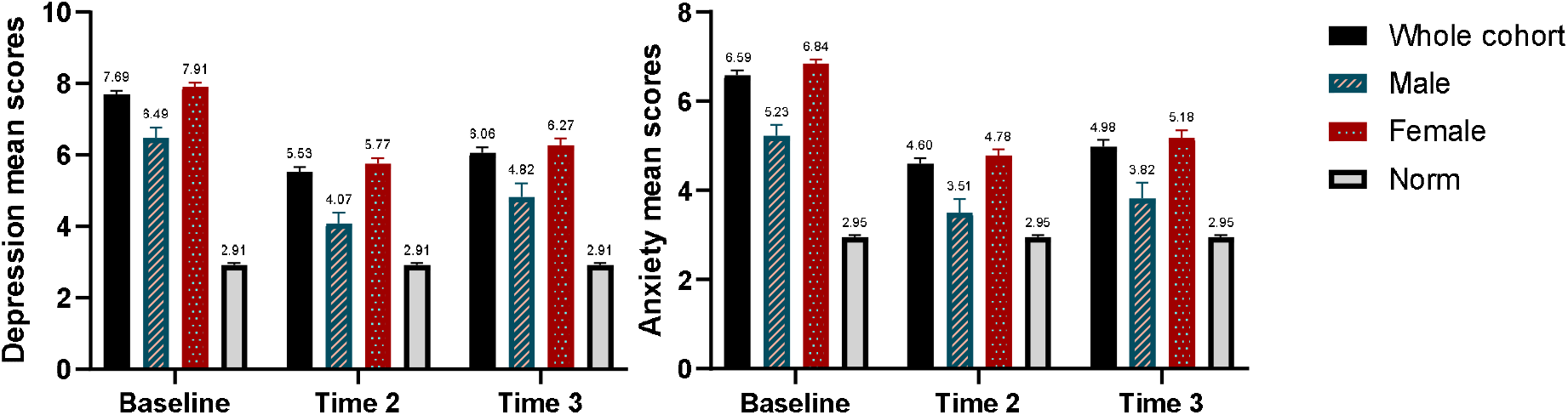
Mean scores with standard errors for depression and anxiety at all three time periods with comparison to population normative data. Bars are mean scores at baseline (n=3097), Time 2 (n=1384) and Time 3 (n=1084). Error bars are standard errors.

When comparing levels of depression and anxiety among completers, significant improvements were seen at Time 2 (depression: *p*<.001; anxiety: *p*<.001). Specifically, mean depression and anxiety scores (square-root transformed) were highest at baseline compared with Time 2 (both *p*<.001) and Time 3 (depression: *p*=.002; anxiety: *p*<.001), while levels at Time 2 were not significantly different from Time 3 (depression: *p*=.10, anxiety: *p*=.054). Similarly, the cases of depression and anxiety according to the original cut-offs(13, 14) showed that fewer completers reported symptoms of depression (49%) and anxiety (37%) at Time 2, compared with baseline and Time 3 (Table 2). A similar pattern was found for ‘caseness’ of depression and anxiety (Table 2). The prevalence of depression and anxiety were 21% and 24% respectively at baseline. At Time 2, the prevalence of depression was 17% which was significantly lower than baseline (21%, *p*=.005) but not Time 3 (19%, *p*=.086). The prevalence of anxiety at Time 2 was 19%, significantly lower than both baseline (24%, *p*=.001) and Time 3 (22%, *p*=.048).

**Table 2:** Categories and cases of depression and anxiety among completers.

|  |  | Baseline | Time 2 | Time 3 |
| --- | --- | --- | --- | --- |
|  |  | n, % | n, % | n, % |
| **Depression (PHQ-9<sup>a</sup>)** |  |  |  |  |
| **Categories** | No-Minimal Depression (0-4) | 428 (48.8) | 470 (53.5) | 440 (50.1) |
|  | Mild Depression (5-9) | 267 (30.4) | 260 (29.6) | 270 (30.8) |
|  | Moderate Depression (10-14) | 119 (13.6) | 97 (11.1) | 98 (11.2) |
|  | Moderately Severe Depression (15-19) | 43 (4.9) | 34 (3.9) | 47 (5.4) |
|  | Severe Depression (20-27) | 21 (2.4) | 17 (1.9) | 23 (2.6) |
| **Cases<sup>b</sup>** | Non-cases (0-9) | 695 (79.2) | 730 (83.1) | 710 (80.9) |
|  | Cases (10-27) | 183 (20.8) | 148 (16.9) | 168 (19.1) |
| **Case incidence and improvement<sup>c</sup>** | *Incidence* | N/A | 59 (6.7) | 48 (5.5) |
|  | *Improvement* | N/A | 94 (10.7) | 21 (2.4) |
| **Anxiety (GAD-7<sup>b</sup>)** |  |  |  |  |
| **Categories** | No-Minimal Anxiety (0-4) | 493 (56.2) | 552 (62.9) | 510 (58.1) |
|  | Mild Anxiety (5-9) | 239 (27.2) | 216 (24.6) | 223 (25.4) |
|  | Moderate Anxiety (10-14) | 78 (8.9) | 68 (7.7) | 91 (10.4) |
|  | Severe Anxiety (15-21) | 68 (7.7) | 42 (4.8) | 54 (6.2) |
| **Cases<sup>b</sup>** | Non-cases (0-7) | 671 (76.4) | 710 (80.9) | 688 (78.4) |
|  | Cases (8-21) | 207 (23.6) | 168 (19.1) | 190 (21.6) |
| **Case incidence and improvement<sup>c</sup>** | *Incidence* | N/A | 53 (6.0) | 45 (5.1) |
|  | *Improvement* | N/A | 92 (10.5) | 24 (2.7) |

<sup>a</sup> PHQ-9, the 9-item Patient Health Questionnaire;<sup>19</sup> GAD-7, the 7-item Generalized Anxiety Disorder Scale.<sup>20</sup>
<sup>b</sup> A 'case' is defined as the PHQ-9 score greater or equal to 10, or the GAD-7 score greater or equal to 8, at which level someone would qualify for high intensity psychological support in the National Health Service
<sup>c</sup> An 'incidence' is defined as becoming a 'case' at Time 2 or 3, 'Improvement' is defined as becoming a 'non-case' at Time 2 or 3.

### Examining risk factors for depression and anxiety

Multivariable linear regression models were constructed to identify prospective significant socio-demographic and baseline psychological predictors of depression and anxiety scores at Time 3 (Table 3). Results showed that being younger (depression: B=-0.14, 95% CI: -0.20, -0.09; anxiety: B=-0.15, 95% CI: -0.21, -0.09 both per 10 year increase), having prior mental health disorders (depression: B=0.56, 95% CI: 0.41, 0.72; anxiety: B=0.51, 95% CI: 0.35, 0.68), and experiencing more negative life events (depression: B=0.24, 95% CI: 0.16, 0.32; anxiety: B=0.19, 95% CI: 0.11, 0.28) were independently and significantly associated with greater depression and anxiety scores at Time 3. In addition, living alone (B=-0.44, 95% CI: -0.67, -0.22) was negatively and significantly associated with greater anxiety. The socio-demographic predictors accounted for 23-24% of the variance (Supplementary Appendix S2). Greater perceived loneliness (depression: B=0.08, 95% CI: 0.04, 0.11; anxiety: B=0.05, 95% CI: 0.02, 0.09) and lower positive mood (depression: B=-0.08, 95% CI:-0.10, - 0.06; anxiety: B=-0.07, 95% CI: -0.09, -0.06) at baseline were independently and significantly associated with both greater depression and anxiety scores at Time 3. Sensitivity analyses with multiply imputed data (n=70) showed that most of these predictors (age, prior mental health disorders, negative life events, loneliness, positive mood) remained significant in models predicting Time 3 depression or anxiety scores (Supplementary Appendix S3). However, in the sensitivity analyses, being female (B=0.17, 95%CI: 0.01, 0.33) and having more COVID-19 worry (most of time, B=0.40, 95%CI: 0.04, 0.75) also significantly predicted greater anxiety at Time 3 (B=0.17, 95%CI: 0.01, 0.33), and experiencing fewer positive life events significantly predicted greater depression (B=-0.20, 95%CI: -0.26, -0.13) and anxiety (B=-0.16, 95%CI: -0.23, -0.09) at Time 3.

**Table 3:** Multivariable linear regression models showing associations between demographic and psychological explanatory variables at baseline and depression and anxiety scores at Time 3.

|  | Depression total score at<br>Time 3<sup>a</sup> | Anxiety total score at<br>Time 3<sup>a</sup> |
| --- | --- | --- |
|  | **B (95% CI), p** | **B (95% CI), p** |
| Age (per 10 year increase) | **-0.14 (-0.20, -0.09), <.001** | **-0.15 (-0.21, -0.09), <.001** |
| Female (yes/no) | 0.04 (-0.16, 0.25), .67 | 0.11 (-0.11, 0.33), .33 |
| BAME background (yes/no) | -0.17 (-0.49, 0.14), .28 | -0.10 (-0.44, 0.23), .55 |
| Key-worker (yes/no) | 0.09 (-0.07, 0.25), .28 | 0.10 (-0.06, 0.27), .22 |
| Prior mental health disorder (yes/no) | **0.56 (0.41, 0.72), <.001** | **0.51 (0.35, 0.68), <.001** |
| Risk Group<sup>b</sup> |  |  |
| Most at Risk | 0.25 (-0.18, 0.68), .26 | 0.11 (-0.35, 0.56), .64 |
| Increased Risk | 0.14 (-0.04, 0.33), .13 | 0.07 (-0.13, 0.27), .47 |
| Living alone (yes/no) | -0.16 (-0.37, 0.06), .15 | **-0.44 (-0.67, -0.22), <.001** |
| Positive life event (per unit) | -0.01 (-0.12, 0.10), .88 | -0.00 (-0.12, 0.12), .95 |
| Negative life event (per unit) | **0.24 (0.16, 0.32), <.001** | **0.19 (0.11, 0.28), <.001** |
| Perceived loneliness (per unit) | **0.08 (0.04, 0.11), <.001** | **0.05 (0.02, 0.09), .005** |
| Positive mood (per unit) | **-0.08 (-0.10, 0.06), <.001** | **-0.07 (-0.09, -0.06), <.001** |
| Perceived risk of COVID-19 (per unit) | 0.01 (-0.02, 0.05), .47 | 0.01 (-0.03, 0.05), .59 |
| COVID-19 worry<sup>c</sup> |  |  |
| No worry | -0.05 (-0.25, 0.15), .63 | -0.11 (-0.33, 0.10), .30 |
| Much of time | 0.10 (-0.14, 0.33), .42 | 0.25 (-0.00, 0.50), .05 |
| Most of time | -0.10 (-0.63, 0.43), .70 | 0.16 (-0.40, 0.72), .58 |
| **Adjusted R<sup>2</sup>** | **0.39** | **0.33** |
| **N** | **717** | **717** |

<sup>a</sup> A square-root transformation was applied to the dependent variable.
<sup>b</sup> Comparison reference group "I am in neither risk category".
<sup>c</sup> Comparison reference group "I occasionally worry about getting COVID-19".

### Cases of depression and anxiety: predictors of change over time

We next distinguished between those who became incident cases of depression and anxiety (i.e., did not meet the criterion for high intensity support at baseline, but did so at either Time 2 or 3) and those who improved (i.e., met criterion for high intensity support at baseline, but were non-cases at Time 2 or 3). At follow-up, 107 (12%) people who were non-cases at baseline became incident depression cases and 98 (11%) became incident anxiety cases. Compared with those who remained a non-case of depression (n=588) or anxiety (n=573) at all times, having prior mental health disorder (depression: OR=3.17, 95% CI: 1.83, 5.47; anxiety: OR=3.93, 95%CI: 2.30, 6.72), experiencing more negative life events (depression: OR=1.35, 95%CI: 1.04, 1.76; anxiety: OR=1.50, 95%CI: 1.17, 1.92), and lower baseline positive mood (depression: OR=0.90, 95% CI: 0.84, 0.96; anxiety: OR=0.92, 95%CI: 0.86, 0.98) were significant independent risk factors for the incidence of depression and anxiety caseness. Greater baseline loneliness was only significantly associated with higher risk of incident depression cases (OR=1.31, 95%CI: 1.16, 1.49) while being younger (OR=0.78, 95%CI: 0.64, 0.94) per 10 years and living with others (OR=0.38, 95%CI: 0.15, 0.94) were only significantly associated with increased risk of incident anxiety cases (Table 4).

**Table 4:** Logistic regression models showing associations between explanatory variables and incidence or improvement of depression and anxiety cases^a^.

|  | Incident depression cases<sup>b</sup> | Incident anxiety cases<sup>b</sup> | Improved depression cases<sup>c</sup> | Improved anxiety cases<sup>c</sup> |
| --- | --- | --- | --- | --- |
|  | **Odd Ratio (95% CI), *p*** | **Odd Ratio (95% CI), *p*** | **Odd Ratio (95% CI), *p*** | **Odd Ratio (95% CI), *p*** |
| Age (per decade) | 1.06 (0.87, 1.29), .57 | **0.78 (0.64, 0.94), .009** | 1.12 (0.85, 1.48), .42 | 1.01 (0.79, 1.28), .96 |
| Prior mental health disorder (yes/no) | **3.17 (1.83, 5.47), <.001** | **3.93 (2.30, 6.72), <.001** | 0.78 (0.39, 1.58), .49 | 0.66 (0.35, 1.27), .22 |
| Live alone (yes/no) | 0.54 (0.25, 1.18), .13 | **0.38 (0.15, 0.94), .04** | 1.44 (0.57, 3.68), .44 | **3.48 (1.31, 9.23), .012** |
| Negative life events (per unit) | **1.35 (1.04, 1.76), .02** | **1.50 (1.17, 1.92), .001** | **0.66 (0.47, 0.91), .011** | 0.76 (0.54, 1.07), .12 |
| Baseline perceived loneliness (per unit) | **1.31 (1.16, 1.49), <.001** | 1.10 (0.97, 1.25), .12 | 0.92 (0.81, 1.05), .22 | **0.84 (0.74, 0.96), .008** |
| Baseline positive mood (per unit) | **0.90 (0.84, 0.96), .003** | **0.92 (0.86, 0.98), .008** | 1.05 (0.96, 1.15), .26 | 1.03 (0.94, 1.12), .54 |
| **Pseudo R<sup>2</sup>** | **0.17** | **0.15** | **0.06** | **0.07** |
| **N** | **652** | **638** | **152** | **177** |

\*\*\* $p < 0.001$ , \*\* $p < 0.01$ , \* $p < 0.05$
<sup>a</sup> A 'case' is defined as the PHQ-9 score greater or equal to 10 for depression, or the GAD-7 score greater or equal to 8 for anxiety, at which level someone would qualify for high intensity psychological support in the National Health Service.
<sup>b</sup> Incidence refers to individuals who were 'non-cases' at baseline and subsequently became 'cases' at Time 2 or 3. The comparison groups were non-cases of depression at all time and non-cases of anxiety at all 3 time, respectively.
<sup>c</sup> Improvement refers to individuals who were 'cases' at baseline and subsequently became 'non-cases' at Time 2 or 3. The comparison groups were non-cases of depression at all time and non-cases of anxiety at all time, respectively.

There were 115 people (13%) who were depression cases at baseline and 116 (13%) who were anxiety cases who improved during follow-up (Table 2). Compared with those who remained a case of depression (n=68) or anxiety (n=91) at all time points, experiencing fewer negative life events (OR=0.66, 95%CI: 0.47, 0.91) was a significant predictor of improved depression cases. Living alone (OR=3.48, 95%CI: 1.31, 9.23) but less loneliness (OR=0.84, 95%CI: 0.74, 0.96) were significant predictors of improved anxiety cases (Table 4).

## Discussion

We report findings from a prospective cohort study established early in the COVID-19 pandemic in the UK longitudinally over the course of 2020.

The overall levels of depression and anxiety showed that both depression and anxiety significantly exceeded pre-pandemic population norms at all three time points, with female participants reporting higher levels of depression and anxiety than male participants. The prevalence of depression and anxiety cases where high intensity psychological support is required in our cohort is worrisome: an overall 17%-21% for depression and 19-24% for anxiety cases. The Office of National Statistics reported that depression made up 14% of all GP diagnoses in 2019.(21) We observed percentages higher than these. Although significant improvements in depression and anxiety were evident after easing of restrictions compared with baseline during the lockdown among completers, such results might only be representative of those who participated in all surveys but not those who dropped out (or survivorship bias).(22) In fact, the levels of depression and anxiety may be underestimated in our cohort, given those with highest levels of depression and anxiety at baseline were less likely to complete further questionnaires. To address this issue, we estimated means and prevalence of depression and anxiety at both Time 2 and 3 using MI data. Estimated mean values for depression (Time 2: 6.44 (SD=0.13), Time 3: 6.83 (SD=0.20)) and anxiety (Time 2: 5.42 (SD=0.13), Time 3: 5.80 (SD=0.16)) were observingly higher than the cross-sectional mean values reported in main analysis (see Figure 1). Estimated prevalence for depression (24%-27%) and anxiety (22%-26%) during follow-ups was also higher than reported in main analysis. These findings together demonstrate the profound disruptions to mental health due to the COVID-19 pandemic and subsequent social restrictions, which may not only be prolonged but may also further influence the physical health of the population.(2)

When looking at what socio-demographic factors were predictive of depression and anxiety later in the pandemic, we found that age continued to be the most important demographic predictor as reported early in the pandemic.(1) And as shown by others, having prior mental health disorders was also a significant predictor.(2, 3) Further, among individuals who were non-cases at baseline, those with prior mental health disorders, were 3-4 times more likely to develop depression or anxiety compared with those without. Experiencing negative life events was another significant risk factor for greater levels of depression and anxiety at Time 3, and of incident depression and anxiety cases. Furthermore, experiencing fewer negative life events was the only significant predictor of improved depression cases. This demonstrates how the direct impacts of COVID-19 on life situations (e.g., loss of close others, changes in employment, financial situations and relationships) can meaningfully impacting mental health among the population. These risk factors of poor mental health were seen in evidence from both before-(23, 24) and during-COVID-19.(4) These results firstly suggest that the demand for mental healthcare might increase further post-restrictions, posing challenges for psychiatry and primary care.(25, 26) Urgent efforts are needed to investigate how to deliver adequate support to those who are in need, and how to prevent deterioration of mental health.(25, 26) Secondly, strategies to cope with the impact of life events, such as bereavement support and wider strategies to improve economy and employment, are also approaches to aid mental health recovery post-pandemic.(27, 28)

When exploring modifiable psychological risk factors of depression and anxiety, we found that greater loneliness and lower positive mood at baseline, significantly predicted depression and anxiety scores in November/December 2020 as seen cross-sectionally in April,(1) after controlling for socio-demographic factors. This is consistent with other evidence.(10, 29) Greater baseline loneliness was a significant risk factor for depression incidence. Lower baseline loneliness, on the other hand, was a significant predictor of improved anxiety. We also found a 10% decrease in the odds of depression or anxiety incidence during follow-up, with per unit of increase in baseline positive mood. This was despite the absence of a relationship between positive mood and improved depression or anxiety. These findings revealed the potential effects of improving social support (and in so doing, reducing loneliness) and positive mood on reducing the risks of depression and anxiety. Positive psychological interventions, featuring elements such as mindfulness, gratitude, and ‘best possible self’ could be among the armoury of approaches we take to address both loneliness and positive mood.(30, 31) However, a range of other approaches are available, such as enhancing social skills and social support, relaxations, and creative activities, all of which have been shown to improve these outcomes.(30, 31)

Some limitations of this work are worthy of note. First, a significant number of participants (72%) dropped out throughout the survey period. This is comparable to other cohorts established early in the pandemic.(22, 32) Those who dropped out were also significantly different compared with those who completed all three surveys, both in demographic characteristics and baseline mental health. The high proportion of drop-outs is likely to have led to an under-estimation of depression and anxiety in our cohort. This was supported by the higher estimated means and prevalence of depression and anxiety from multiple imputations. Indeed, reaching and retaining individuals most in need of mental health support is not uncommon in such research.(22) The high proportion of keyworkers in our cohort (50% at baseline) may have also contributed to the high drop-out rate. These individuals were, by definition, providing essential services throughout the pandemic and will, therefore, have had less capacity to remain engaged in the research. However, it is unlikely that the socio-demographic and psychological predictors of anxiety and depression identified in our models were affected by drop-outs. In the multivariable regression models with MI data, most of the predictors of Time 3 depression and anxiety (i.e., age, prior mental health disorders, negative life events, loneliness, positive mood) were still significant. Another limitation includes the absence of health behaviour data (e.g., on physical activity, sleep quality etc.) which may also have contributed to mental health outcomes.(9, 33)

## Conclusion

Our findings indicate that the effects of the COVID-19 pandemic on mental health have been profound and persisted throughout 2020. Despite modest improvements with the easing of restrictions, levels of anxiety and depression remained stubbornly higher than pre-pandemic levels. Consistent with previous work, being female, younger age and having a previous history of mental health difficulties were associated with a greater risk of anxiety and depression. However, our findings on modifiable predictors (i.e., loneliness and positive mood) highlight potential avenues for intervention.

## Data Availability

Data will be deposited in the University of Nottingham data archive. Access to this dataset will be embargoed for a period of 12 months to permit planned analyses of the dataset. After that it may be shared with the consent of the Chief Investigator. Extra data is available by contacting.

## Declaration of Interest

All authors have completed the ICMJE uniform disclosure form at www.icmje.org/coi_disclosure.pdf and declare: no support from any organisation for the submitted work; no financial relationships with any organisations that might have an interest in the submitted work in the previous three years; no other relationships or activities that could appear to have influenced the submitted work.

## Funding

KA acknowledges the financial support from the National Institute for Health Research School for Primary Care Research (NIHR SPCR). TC acknowledges the financial support of the Department of Health via the National Institute for Health Research (NIHR) Specialist Biomedical Research Centre for Mental Health award to the South London and Maudsley NHS Foundation Trust (SLaM) and the Institute of Psychiatry at King’s College London. CC acknowledges support from the NIHR Nottingham Biomedical Research Centre. The views expressed are those of the authors and not necessarily those of the NHS, the NIHR or the Department of Health and Social Care. No other funding supported the work described in this manuscript.

## Acknowledgements

We would like to thank all our participants and the NHS organisations who helped us promote the study.

## Author Contribution

All authors contributed to the study design. RJ led statistical analyses alongside KA and CC. RJ led the drafting of the manuscript but all authors contributed to the preparation and review of the final manuscript and approved for submission.

## Notes

### Competing Interest Statement

The authors have declared no competing interest.

### Author Declarations

The University of Nottingham Faculty of Medicine and Health Sciences (ref: 506-2003) and the NHS Health Research Authority (ref: 20/HRA/1858) approved all study procedures.

