## Supplementary material for "The prevalence, incidence, prognosis and risk factors for depression and anxiety in a UK cohort during the COVID-19 pandemic": Supplimentary appendix

**Supplementary Appendices**

**Appendix S1: Measures for prior mental health disorder and positive/negative life events**

**Table 1 Question items for prior mental health disorder and positive/negative life events**

|  | **Question** | **Response options** | **Code** |
| --- | --- | --- | --- |
| **Prior mental health disorder** | Have you had the experience of being diagnosed with any mental health issues (e.g., depression/anxiety/PTSD) previously? | Yes/No | Prior mental health disorder (Yes/No) |
| **Experience of life events** | Due to the COVID-19 outbreak, have the following events happened at some point? (Please select all that apply) | Death of a spouse/ partner/ close relative or friend | Negative |
|  |  | Major health event for you or a loved one requiring hospitalisation | Negative |
|  |  | You or your partner losing your job | Negative |
|  |  | Gaining new employment | Positive |
|  |  | Change in financial status for the better (e.g. earning more money) | Positive |
|  |  | Change in financial status for the worse (e.g. hours of employment reduced) | Negative |
|  |  | Change in living conditions for the better | Positive |
|  |  | Change in living conditions for the worse | Negative |
|  |  | Change in personal relations for the better | Positive |
|  |  | Change in personal relations for the worse | Negative |
|  |  | None of the above | N/A |

**Appendix S2: Results from univariable regressions**

**Table 2 Regression models showing associations between demographic explanatory variables and depression and anxiety scores at Time 3**

|  | **Depression total score at Time 3^a^** | **Anxiety total score at Time 3 ^a^** |
| --- | --- | --- |
|  | **B (95%CI), *p*** | **B (95%CI), *p*** |
| Age (per decade) | -0.21(-0.26, -0.15), <.001 | -0.23 (-0.28, -0.17), <.001 |
| Female (yes/no) | 0.14 (-0.07, 0.34), .20 | 0.15 (-0.07, 0.37), .17 |
| BAME background (yes/no) | 0.03 (-0.28, 0.35), .83 | 0.01 (-0.32, 0.34), .94 |
| Key-worker (yes/no) | 0.11 (-0.04, 0.26), .14 | 0.16 (0.01, 0.32), .040 |
| Prior mental health disorder (yes/no) | 0.72 (0.57, 0.87), <.001 | 0.73 (0.58, 0.89), <.001 |
| Risk Group ^b^ |  |  |
| Most at Risk | 0.55 (0.12, 0.99), .014 | 0.50 (0.04, 0.95), .032 |
| Increased Risk | 0.15 (-0.03, 0.34), .10 | 0.16 (-0.03, 0.35), .10 |
| Living alone (yes/no) | 0.15 (-0.06, 0.35), .16 | -0.17 (-0.39, 0.04), .11 |
| Positive life event (per unit) | -0.09 (-0.20, 0.01), .089 | -0.13 (-0.24, -0.02), .026 |
| Negative life event (per unit) | 0.31 (0.23, 0.39), <.001 | 0.25 (0.17, 0.34), <.001 |
| **Adjust R^2^** | **0.24** | **0.23** |
| **N** | **869** | **869** |

^a^ A square-root transformation was applied to the dependent variable.

^b^ Comparison reference group “I am in neither risk category”.

**Appendix S3: Sensitivity analysis**

**Table 3 Regression models showing associations between sociodemographic and psychological explanatory variables and depression or anxiety scores at Time 3 with 70 imputed datasets**

|  | **Depression total score at Time 3^a^** | **Anxiety total score at Time 3^a^** |
| --- | --- | --- |
|  | **B (95%CI), *p*** | **B (95%CI), *p*** |
| Age (per decade) | -0.15 (-0.19, -0.11), <.001 | -0.17 (-0.22, -0.13), <.001 |
| Female (yes/no) | 0.14 (-0.01, 0.29), .071 | 0.17 (0.01, 0.33), .040 |
| BAME background (yes/no) | 0.03 (-0.19, 0.25), .76 | 0.01 (-0.22, 0.23), .96 |
| Key-worker (yes/no) | 0.01 (-0.11, 0.13), .89 | 0.05 (-0.06, 0.15), .39 |
| Prior mental health disorder (yes/no) | 0.57 (0.44, 0.70), <.001 | 0.61 (0.47, 0.75), <.001 |
| Risk Group ^b^ |  |  |
| Most at Risk | 0.22 (-0.09, 0.53), .17 | 0.03 (-0.29, 0.36), .85 |
| Increased Risk | 0.06 (-0.09, 0.21), .40 | 0.09 (-0.07, 0.25), .29 |
| Living alone (yes/no) | -0.04 (-0.20, 0.12), .66 | -0.25 (-0.42, -0.09), .003 |
| Positive life event (per unit) | -0.20 (-0.26, -0.13), <.001 | -0.16 (-0.23, -0.09), <.001 |
| Negative life event (per unit) | 0.13 (0.09, 0.18), <.001 | 0.11 (0.06, 0.16), <.001 |
| Perceived loneliness (per unit) | 0.08 (0.05, 0.11), <.001 | 0.08 (0.05, 0.11), <.001 |
| Positive mood (per unit) | -0.07 (-0.09, -0.06), <.001 | -0.06 (-0.08, -0.05), <.001 |
| COVID-19 worry ^c^ |  |  |
| No worry | -0.03 (-0.18, 0.12), .68 | -0.16 (-0.33, 0.01), .067 |
| Much of time | -0.01 (-0.18, 0.16), .92 | 0.12 (-0.07, 0.30), .21 |
| Most of time | 0.15 (-0.19, 0.49), .40 | 0.40 (0.04, 0.75), .027 |
| **N** | **3094** | **3094** |

^a^ A square-root transformation was applied to the dependent variable.

^b^ Comparison reference group “I am in neither risk category”.

^c^ Comparison reference group “I occasionally worry about getting COVID-19”.
